# Chronic psychosocial stress is associated with higher MRI-visible perivascular space volumes in healthy young adults

**DOI:** 10.64898/2026.04.29.26352039

**Authors:** Jose Bernal, Igor Izyurov, Marina Krylova, Nathalie Winter, Maria del C. Valdés-Hernández, Roberto Duarte Coello, Joanna M. Wardlaw, Soroosh Golbabaei, Luisa Herrmann, Louise Martens, Daniel Güllmar, Laith Hamid, Anja Buder, Marc Dörner, Katja Neumann, Hendrik Mattern, Veronika Engert, Stefanie Schreiber, Martin Walter, Lejla Colic

## Abstract

Chronic psychosocial stress (CPS) is associated with adverse brain and mental health outcomes. Effects on the cerebral microvasculature have been proposed as an underlying mechanism, although this remains to be established. Here, we examined the association between CPS and an early marker of microvascular dysfunction, magnetic resonance imaging (MRI)-visible perivascular spaces (PVS). Analyses were conducted in two cohorts of healthy young adults (N = 61; ages 18-43 years; 88% male) using high-resolution 3T MRI and an automated PVS quantification pipeline. CPS was assessed using the Perceived Stress Scale (PSS-10). We applied a two-step meta-analytic framework and controlled for known allostatic factors impacting PVS, including age, body mass index and mean arterial pressure. In accordance with our hypothesis, individuals with higher CPS had significantly higher fractional PVS volumes in the centrum semiovale (CSO), in particular in the frontal and occipital lobes (p_FDR_ < .05). No such effect was found in the basal ganglia, or in the CSO subdivision, parietal, and temporal lobes (p_FDR_ > .09). Our findings indicate that CPS may contribute to subtle, centrum semiovale specific microvascular alterations even in healthy young adults. Future multimodal research including inflammatory marker and blood-brain barrier measures may help to elucidate mechanistic pathways.

## Introduction

Chronic psychosocial stress (CPS) is increasingly recognized as a contributor to poor mental and physical health outcomes (Agorastos and Chrousos 2022), potentially by creating allostatic load on physiological systems (Guidi et al. 2020). Allostatic load arises from the brain and body’s interpretations and reactions to stressful stimuli, which can lead to maladaptive changes in brain and body physiology (McEwen 2016).

CPS has effects on brain health, with downstream implications for mental health. Epidemiological and occupational studies have shown that CPS is associated with higher symptom levels of depression, anxiety and burnout (Siegrist 2008, Agyapong et al. 2022). CPS also relates to gross structural brain measures, such as reduced brain volume (Blix et al. 2013) or cortical thickness (Cardoner et al. 2024, Li et al. 2025). The primary mechanisms proposed to link CPS, brain alterations, and mental health include neuroendocrine (Coffman 2020), microvascular (Liu et al. 2017, Mak et al. 2023) and inflammatory pathways (Rohleder 2019), which are likely interrelated and influence each other. Brain microvasculature pathways are particularly relevant, as their dysfunction has well-documented effects on the development of mental health conditions (Clancy et al. 2021, van Agtmaal et al. 2017) and dementia disorders (Brain et al. 2023, Bos et al. 2018).

A promising non-invasive strategy for detecting early cerebral microvascular dysfunction is the quantification of perivascular spaces (PVS) via magnetic resonance imaging (MRI) (Francis, Ballerini, and Wardlaw 2019, Okar et al. 2023, Wardlaw et al. 2020). PVS are a brain-wide network of microscopic channels around blood vessels that facilitate cerebrospinal fluid circulation (Cengija, Eide, and Ringstad 2026, Hirschler et al. 2025, Yamamoto et al. 2024) and support the clearance of metabolic and neurotoxic waste (Braun and Iliff 2020, Hablitz and Nedergaard 2021). Despite their regular microscopic size, these spaces can enlarge, sometimes reaching a size that makes them visible *in vivo* on MRI (Wuerfel et al. 2008, Barisano et al. 2025). Such enlargements are pathological (Bown et al. 2022, Francis, Ballerini, and Wardlaw 2019) and associate with neurological conditions such as cerebral small vessel disease (Charidimou et al. 2022), allostatic markers such as ageing (Lynch et al. 2022, Menze et al. 2024, Kim et al. 2022), hypertension (Zhu et al. 2010, Jiménez-Balado et al. 2018), metabolic syndrome symptoms (Qi et al. 2021, Cai et al. 2022), sleep deprivation and disorders (Sommer et al. 2025, Einspänner et al. 2025), as well as worse cognitive performance and dementia (Chappell et al. 2025, Barisano et al. 2025). Thus, investigating PVS in relation to CPS may help elucidate the pathophysiological link between chronic stress and microvascular dysfunction.

Here, we leveraged two samples of healthy young participants who underwent high resolution T1-weighted imaging at 3T and completed self-reported questionnaire on CPS. We used an automated pipeline to segment PVS and applied a meta-regression approach, controlling for known allostatic factors such as age, body mass index and mean arterial pressure. We hypothesized that higher levels of CPS would be associated with increased PVS volumes.

## Methods

### Participants

Study 1 (site Jena) was designed to investigate the effects of childhood adversity on acute stress response using simultaneous electroencephalography (EEG) and magnetic resonance imaging (MRI). In addition to structural MRI, the study included functional MRI (fMRI) and magnetic resonance spectroscopy (MRS), which together with EEG were not analyzed in the current study. This study was approved by the Ethical committee of Jena University Hospital (2022-2718-1-BO). All participants gave written informed consent and were reimbursed for their participation. Study inclusion criteria were an age between eighteen to fifty-fives years, and fluency in German. Both sexes were included. Exclusion criteria were lifetime psychotic disorder/ symptoms or any psychiatric disorder within the past year according to Diagnostic and Statistical Manual of Mental Disorders (DSM)-5 criteria (American Psychiatric Association and American Psychiatric 2013), use of psychotropic medications within past year, instable somatic disease (e.g., untreated thyroid disorder), left-handedness and any other MRI exclusion criteria. The study was run between 11/2022 until 05/2023. In total, twenty-seven participants were recruited. One participant did not wish to continue the study after the initial visit, one was excluded based on the clinical interview, and three were not scanned due to technical difficulties with the scanner. One participant had high motion during the T1-weighted scan and two did not have information on the diastolic blood pressure. Thus, nineteen participants were included in the current analysis (age range 18-43 years, mean age 27.5, standard deviation [SD; 8.1], 12 [63%] men).

Study 2 (site Tübingen) was designed to investigate effects of childhood adversity and genetic polymorphism in the GAD65 gene on stress markers. In addition to structural MRI, the study included fMRI and MRS that were not analyzed in the current study. The study was approved by the Ethical committee of Medical Faculty of Eberhard Karls University Tübingen (578/2016B01). All participants gave written informed consent and were reimbursed for their participation. Study inclusion criteria were male sex, ages between eighteen to forty years, non-smoking, and fluency in German. Exclusion criteria were a history of psychiatric disorders determined by the Structured Clinical Interview (Wittchen et al. 1997) according to the DSM-IV criteria (Guze 1995), instable somatic disease, substance abuse in the past three months (including nicotine) and any MRI exclusion criteria (e.g., neurological conditions, implants). In total, fifty-eight participants were recruited. The study was run between 12/2017 until 07/2019. Eleven participants did not wish to continue the study after the initial visit; three participants could not be scheduled while one participant did not fill out questionnaires. Thus, forty-two participants were included in the current analysis (age range 20-32 years, mean age 25.9, SD [3.3]).

### Psychometric assessment

The German version of the PSS-10 (Klein et al. 2016) was used to evaluate chronic psychosocial stress in both studies. PSS-10 items focus on the overall perception of stress load rather than specific events (Cohen, Kamarck, and Mermelstein 1994) and the questionnaire is used as a measure for chronic psychosocial stress (Patterson, Sagui-Henson, and Prather 2020). Responses are given on a Likert scale ranging from 0-4. In Study 1, Cronbach’s raw alpha was 0.79, while in Study 2 it was 0.80, indicating good reliability.

### Magnetic resonance imaging

In both studies structural MRI data were acquired using a Siemens 3T Prisma scanner (Siemens Healthineers, Erlangen, Germany) equipped with a 64-channel head coil. Acquisition parameters differed between the studies.

In study 1 (site Jena) high-resolution T1-weighted Magnetization Prepared Rapid Acquisition with Gradient Echoes (MPRAGE) images were acquired with: sagittal orientation, acceleration mode GRAPPA, factor PE= 2, FOV= 240 × 256 × 167 mm, bandwidth= 220 Hz/Px, slices= 208, TR= 2400 ms, TE= 2.22 ms, flip angle= 8°, and voxel size= 0.8 mm^3^ isotropic, acquisition time= 6:38 min.

In study 2 (site Tübingen) high-resolution T1-MPRAGE images were acquired with: no imaging acceleration, factor PE= 1, field of view (FOV)= 256 × 256 × 192 mm, bandwidth= 150 Hz/Px, slices= 192, repetition time (TR)= 2300 ms, echo time (TE)= 4 ms, flip angle= 9°, and voxel size= 1 mm^3^ isotropic, acquisition time= 6:20 min.

### Region of interest segmentation

We applied SynthSeg (Billot et al. 2023) to the T1-weighted images to obtain subject-specific whole-brain parcellations. These parcellations served as the basis for deriving anatomically defined masks of the basal ganglia and centrum semiovale regions of interest (BG ROI and CSO ROI). The BG ROI included the internal and external capsules, caudate nucleus, lentiform nucleus, and thalamus, whereas the CSO ROI comprised the remaining supratentorial white matter. Owing to the large extent and anatomical heterogeneity of the CSO ROI, we further subdivided this region using the USCBrainLobes Atlas (Joshi et al. 2022) to generate lobar masks (frontal, occipital, temporal, and parietal), thereby enabling a more spatially resolved and anatomically specific regional analysis. The USCBrainLobes Atlas was registered to each individual case using SynthMorph (Hoffmann et al. 2021) via a joint affine and deformable transformation, after which the atlas label map was propagated to individual subject space using nearest-neighbor interpolation.

### PVS segmentation and measurements

We quantified PVS from T1-weighted MRI using a previously validated automated segmentation pipeline (Bernal et al. 2025, Bitar et al. 2025, Menze et al. 2024, Hernandez et al. 2024). We performed segmentation with DRIPS, a deep learning-based framework that demonstrates high accuracy and robustness for PVS detection across both T1- and T2-weighted MRI acquisitions (Bitar et al. 2025). All segmentations were visually inspected and manually refined by a trained image analyst (JoBe) to minimize false positives. These corrections primarily addressed occasional misclassifications arising from imperfections in SynthSeg-derived anatomical parcellations, especially within occipital lobes. Once PVS segmentation and manual correction were completed, we estimated regional PVS burden as fractional PVS volume within each of the aforementioned regions of interest (PVS volume within the ROI divided by the total ROI volume).

### Data analysis

We employed a two-step meta-analytic framework to account for substantial differences between cohorts in imaging protocols, population composition, and, consequently, PVS measurement characteristics. We first estimated the association between PSS-10 scores (outcome) and regional fractional PVS volumes (predictor) separately within each cohort using linear regression models adjusted for age, BMI, and mean arterial pressure.

Prior to model fitting, we transformed all variables using the *bestNormalize* package (Peterson and Cavanaugh 2020) in R to address skewness and improve distributional properties. Across both studies, ordered quantile normalization was applied to regional fractional PVS volumes and age, while BMI and mean arterial pressure were transformed using Box–Cox normalization. For PSS scores, a square-root transformation was applied in Study 1, whereas ordered quantile normalization was used in Study 2.

We then combined cohort-specific effect estimates using inverse-variance weighted random-effects meta-analysis with restricted maximum likelihood (REML), as implemented in the *metafor* package in R (Viechtbauer 2010). To account for multiple comparisons, false discovery rate (FDR) correction was applied using the Benjamini-Hochberg procedure, controlling the expected proportion of false positives among statistically significant findings. Effects were considered significant at p_FDR_ < .05.

## Results

### Study characteristics

A total of 61 individuals were included in the analysis (19 from Study 1 and 42 from Study 2). The clinical and imaging characteristics of both cohorts are summarized in **Table 1**. The two studies differed in terms of sex distribution, mean arterial pressure, PSS-10 scores, and PVS volumes within the CSO region of interest, specifically in the frontal, parietal, and occipital lobes. This variability across numerous parameters strongly supports the use of a two-step meta-analytic framework rather than a conventional pooled linear regression with interaction terms, as many variables of interest and potential confounders differ substantially between studies.

**Table 1.** Clinical and imaging characteristics for each cohort. Values are presented as untransformed, median [min, max] or number of participants [percentage]. Differences in sex distribution between studies were assessed using Pearson’s chi-square test, while continuous variables were compared using the Wilcoxon rank-sum test. The corresponding statistics are shown in the right-most column.

|  | Study 1<br>(N = 19) | Study 2<br>(N = 42) | Statistics |
| --- | --- | --- | --- |
| Age (years) | 23 [18, 43] | 26 [20, 32] | $W = 415, p = .80$ |
| Males (n, %) | 12 (63%) | 42 (100%) | $\chi^2 (1) = 14.04, p < .001$ |
| Body mass index | 23.8 [19.1, 29.4] | 24.4 [18.2, 32.8] | $W = 496, p = .13$ |
| Mean arterial pressure | 92.3 [76.7, 109] | 95.8 [83.7, 124] | $W = 530, p = .042$ |
| PSS-10 scores | 8 [0, 18] | 13 [2, 31] | $W = 638.5, p < .001$ |
| Fractional BG PVS volumes (% of ROI) | 0.27 [0.11, 0.61] | 0.29 [0.08, 0.44] | $W = 436, p = .57$ |
| Fractional CSO PVS volumes (% of ROI) | 0.10 [0.05, 0.34] | 0.04 [0.01, 0.72] | $W = 135, p < .001$ |
| Fractional frontal PVS volumes (% of ROI) | 0.03 [0.001, 0.14] | 0.01 [0.00, 0.22] | $W = 150, p < .001$ |
| Fractional parietal PVS volumes (% of ROI) | 0.07 [0.01, 0.21] | 0.01 [0.00, 0.56] | $W = 124, p < .001$ |
| Fractional occipital PVS volumes (% of ROI) | 0.02 [0.00, 0.08] | 0.004 [0.00, 0.13] | $W = 148.5, p < .001$ |
| Fractional temporal PVS volumes (% of ROI) | 0.05 [0.01, 0.12] | 0.04 [0.001, 0.25] | W = 357, p = .52 |

### PSS-10 scores are associated with PVS measurements

Individuals with higher PSS-10 scores had higher fractional CSO PVS volumes (β = 0.33, 95% CI [0.08, 0.59], p = .011, p_FDR_ = .038), whereas no association was observed with fractional BG PVS volumes (β = −0.09, 95% CI [−0.35, 0.17], p = .484, p_FDR_ = .535). Given the broad anatomical extent of the CSO ROI, we further examined this region at a finer lobar resolution. This revealed that individuals with higher PSS-10 scores also had higher fractional PVS volumes in the occipital (β = 0.34, 95% CI [0.06, 0.61], p = .016, p_FDR_ = .038) and frontal lobes (β = 0.32, 95% CI [0.05, 0.59], p = .019, p_FDR_ = .038). No significant associations were observed for fractional parietal (β = 0.26, 95% CI [0.00, 0.52], p = .048, p_FDR_ = .073) or temporal PVS volumes (β = −0.09, 95% CI [−0.38, 0.20], p = .534, p_FDR_ = .535). No between-study heterogeneity was observed across any of the analyses (I^2^ = 0%).

**Figure 1.**
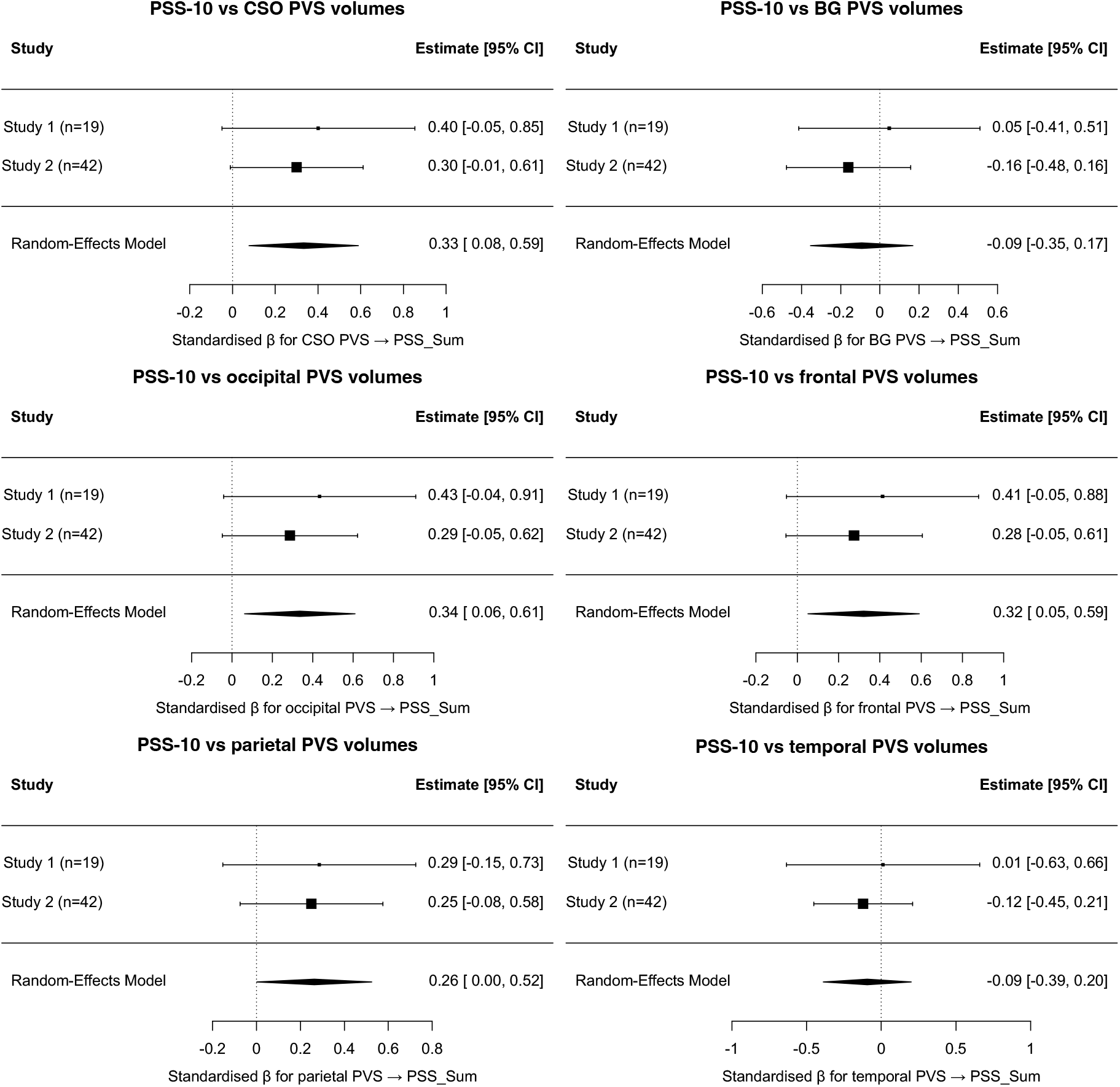
Association between PSS-10 and regional, fractional PVS volumes. Forest plots show cohort-specific and pooled random-effects estimates for the association between PSS-10 scores and fractional PVS volumes, adjusted for participant age, body mass index, and mean arterial pressure. (A) Primary analyses show a significant association between PSS-10 scores and fractional CSO PVS volume, but not fractional BG PVS volume. (B) Secondary lobar analyses of the CSO show significant associations in the frontal and occipital lobes, but not in the parietal or temporal lobes. Squares represent cohort-specific estimates, with size proportional to inversevariance weight, and diamonds represent pooled estimates. Error bars indicate 95% confidence intervals.

## Discussion

CPS imposes a major allostatic burden on the organisms and is linked to adverse health outcomes, including effects on brain health. The underlying mechanisms of this association remain unclear. As microvascular alterations have been proposed as one pathway linking CPS to health-related outcomes, we examined whether CPS is associated with PVS, an imaging marker of early cerebral microvascular dysfunction. We observed that healthy young adults with higher CPS severity had higher fractional PVS volume in the centrum semiovale (CSO) ROI, specifically in frontal and occipital regions. These associations remained significant after adjustment for other allostatic factors, including age, body mass index and mean arterial pressure, suggesting that CPS is indeed associated with early alterations in the cerebral microvasculature. This may be of importance as enlarged PVS may be attenuated with positive lifestyle changes before irreversible tissue damage takes place (Piechowiak et al. 2025).

There is growing interest in understanding whether CPS and microvascular dysfunction may exert synergistic effects that increase vulnerability to worsening brain health. A possible candidate mechanism is the unresolved activation of the immune system, which may act through increased blood brain barrier (BBB) permeability. CPS has indeed been associated with chronic low-grade inflammation (Hänsel et al. 2010, Johnson, Abbasi, and Master 2013). Animal models of chronic social defeat stress also demonstrate higher immune activation (Stewart et al. 2015) as well as BBB dysregulation (Menard et al. 2017).

As integral components of the neurovascular unit, PVS are units that connect neurovascular, clearance, and neuroinflammatory mechanisms (Owens, Bechmann, and Engelhardt 2008, Troili et al. 2020). They are also among the earliest structures affected in cerebral microvascular dysfunction (Waymont et al. 2024, Wardlaw et al. 2020). Higher number and volume of PVS have been also documented as markers of brain psychopathological processes. Cross-sectional studies in humans have shown an association between enlarged PVS and markers of BBB disruption (Li et al. 2019) as well as levels of interleukin 6 (Aribisala et al. 2014). Furthermore, animal models have shown strong relations between chronic stress, BBB disruption and depressive-like symptoms (Matsuno et al. 2022, Welcome 2020). Lastly, initial mechanistic evidence suggests an association between BBB integrity and PVS in an inflammatory animal model (Erickson et al. 2023).

Apart from the immune system, another possible moderator of the link between CPS and brain health may be metabolic symptoms. Even though metabolic symptoms are related to older age, a meta-analysis reported that around 5% of young people have some metabolic syndrome symptoms (Nolan et al. 2017). Metabolic syndrome has been also related to CPS (Pedersen et al. 2016, Kuo et al. 2019). Features of metabolic syndrome such as arterial stiffness (Boutouyrie et al. 2021) and hyperlipidemia (Stokes et al. 2002) contribute to deterioration of small capillaries in the brain potentially contributing to the enlargement of PVS. Recent studies showed that in healthy populations indices of metabolic syndrome were related to PVS (Cai et al. 2022), especially in the CSO (Qi et al. 2021).

Consistent with previous studies (Cengija, Eide, and Ringstad 2026, Arndt et al. 2026, Libecap et al. 2023), our findings suggest spatial specificity, with CPS associated with PVS in the CSO ROI, but not in the BG ROI. Further work is nonetheless needed to pinpoint the mechanisms underlying this regional pattern.

PVS have most often been quantified in relation to cognitive symptoms in ageing (Park et al. 2023, Lynch et al. 2022, Bown et al. 2022) or dementia studies (Menze et al. 2024); for example, higher PVS count has been related with poorer cognition in neurological disorders (Kim et al. 2024, Chappell et al. 2025). Studies examining other psychological factors are less common. Nevertheless, there are few studies that corroborate our findings. Recent work used high-field 7 Tesla MRI to investigate PVS differences between individuals with depression and healthy controls. While no group differences were found, there was a positive association between the number of traumatic events and PVS count, suggesting that stress exposure, rather than depression diagnosis per se, may be linked to increased PVS (Ranti et al. 2022). Another study in individuals with post-COVID syndrome found a positive association between low education attainment and PVS (Bernal et al. 2025). Education is a core element of socio-economic status which has been related with increased chronic stress (Kopp et al. 2007, Baum, Garofalo, and Yali 1999). Moreover, a large population study (>1000 participants) found an association among CSO PVS count and hair cortisol (Hernández et al. 2025), a physiological marker of chronic stress (Wippert et al. 2014). In a mixed cohort of individuals with cognitive impairments and healthy controls, the association between baseline salivary cortisol and PVS was moderated by inflammatory markers (Sibilia et al. 2024), indicating an interplay between HPA-axis, inflammation and brain structure. Together, these studies indirectly support the idea that PVS may reflect very early changes in stress-related brain morphology and may increase risk for later health problems.

This study however has some limitations. First, we investigated two healthy young populations which were mostly males. Women (Matud 2004, Gilbert-Ouimet et al. 2020) and older age groups (particularly those in midlife; (Almeida et al. 2020) tend to experience increased levels of CPS. Thus, future studies should focus on these populations. Second, our study relied on chronic psychosocial stress derived from a self-reported retrospective questionnaire, which may introduce recall bias. New methods like ecological momentary assessment combined with wearable devices that prospectively measure experienced stress and blood pressure could allow for a more detailed examination of the relationship between CPS and brain microvasculature indices. Third, although the T1-weighted scans used were high resolution (0.8 and 1 mm^3^), using even higher resolution imaging, like 7 Tesla MRI, may allow for detection of smaller PVS (Barisano et al. 2020, Einspänner et al. 2025). Finally, we did not measure inflammatory, BBB or metabolic syndrome indicators, which may further moderate or explain the mechanisms linking CPS and PVS.

## Conclusions

The current results suggest a positive association between CPS and PVS volumes in the CSO ROI, particularly within the frontal and occipital lobes, in two healthy samples of young individuals, while controlling for relevant covariates. The findings point to a potential link between CPS and early microvascular alterations in the brain, suggesting one mechanism though which CPS may affect brain health over time. In the longer term, associations between CPS, PVS and inflammation and/or BBB integrity should be investigated using multimodal assessments.

## Data Availability

All data produced in the present study are available upon reasonable request to the authors.

## Authors contributions

JoBe: Methodology; Formal analysis; Writing - Original Draft. IgIz: Investigation; Writing - Review & Editing. MaKry: Investigation; Writing - Review & Editing. NaWi: Investigation; Data Curation; Writing - Review & Editing. MaVal: Methodology; Writing - Review & Editing. RoDuCo: Methodology; Writing - Review & Editing. JoWar: Methodology; Writing - Review & Editing. SoGo: Investigation; Writing - Review & Editing. LuHer: Investigation; Data Curation; Writing - Review & Editing. LoMa: Investigation; Data Curation; Writing - Review & Editing. DaGü: Methodology; Writing - Review & Editing. LaHa: Project administration; Writing - Review & Editing. AnjBu: Project administration; Writing - Review & Editing. MaDör: Methodology; Writing – Review & Editing. KaNeu: Methodology; Writing - Review & Editing. HenMat: Methodology; Writing - Review & Editing. VeEng: Conceptualization; Writing - Review & Editing. SteSch: Conceptualization; Writing - Review & Editing. MaWa: Funding acquisition; Conceptualization; Writing - Review & Editing. LeCo: Conceptualization; Investigation; Data Curation; Supervision; Writing - Original Draft.

## Declaration of generative AI and AI-assisted technologies in the manuscript preparation process

During the preparation of this work the authors used Nature Research Assistant to improve grammar and clarity of paragraphs. After using these tools, the authors reviewed and edited the content as needed and took full responsibility for the content of the published article.

## Acknowledgement

We thank our participants for their participation in our studies. We thank research assistants and staff that participated in the data collection (site Tübingen: Greta Amedick, Vanessa Kasties, Rahel Gutbrod and Mia Meike; site Jena: Emilija Romic, Selina Bunghardt, Marie Emmermann, Elisabeth Proba; and MTRA Ines Krumbein).

